# Combining serological assays and official statistics to describe the trajectory of the COVID-19 pandemic: results from the EPICOVID19-RS study in Rio Grande do Sul (Southern Brazil)

**DOI:** 10.1101/2021.05.21.21257634

**Authors:** Fernando P. Hartwig, Luís Paulo Vidaletti, Aluísio J. D. Barros, Gabriel D. Victora, Ana M. B. Menezes, Marilia A. Mesenburg, Bernardo L. Horta, Mariângela F. Silveira, Cesar G. Victora, Pedro C. Hallal, Claudio J. Struchiner

## Abstract

**Background:** The EPICOVID19-RS study conducted 10 population-based surveys in Rio Grande do Sul (Southern Brazil), starting early in the epidemic. The sensitivity of the rapid point-of-care test used in the first eight surveys has been shown to decrease over time after some phases of the study were concluded. The 9^th^ survey used both the rapid test and an enzyme-linked immunosorbent assay (ELISA) test, which has a higher and stable sensitivity.

**Methods:** We provide a theoretical justification for a correction procedure of the rapid test estimates, assess its performance in a simulated dataset and apply it to empirical data from the EPICOVID19-RS study. COVID-19 deaths from official statistics were used as an indicator of the temporal distribution of the epidemic, under the assumption that fatality is constant over time. Both the indicator and results from the 9^th^ survey were used to calibrate the temporal decay function of the rapid test’s sensitivity from a previous validation study, which was used to estimate the true sensitivity in each survey and adjust the rapid test estimates accordingly.

**Results:** Simulations corroborated the procedure is valid. Corrected seroprevalence estimates were substantially larger than uncorrected estimates, which were substantially smaller than respective estimates from confirmed cases and therefore clearly underestimate the true infection prevalence.

**Conclusion:** Correcting biased estimates requires a combination of data and modelling assumptions. This work illustrates the practical utility of analytical procedures, but also the critical need for good quality, populationally-representative data for tracking the progress of the epidemic and substantiate both projection models and policy making.

## 1. INTRODUCTION

Brazil concentrates 10% of all COVID-19 deaths in the world while having only 2.7% of the world’s population. As of February 28, 2021, Brazil is the country ranking second and third in terms of deaths and confirmed cases, respectively. The situation was worsened the rapid spread of a new variant of the SARS-CoV-2 virus in the Amazon region in the North(1), which is already circulating in other regions of the country, including the South region(2). Rio Grande do Sul (the Southernmost state in Brazil) is facing the worst situation since the start of the pandemic, with infections increasing at unprecedented rates. In February 27 2021, all subregions of the state were, for the first time, classified as being in a critical situation based on a controlled social distancing model that uses several indicators and was launched by the State’s Government in May 2020(3).

Rio Grande do Sul was one of the first regions in the world to have a large population-based assessment of the COVID-19 epidemic. According to official statistics, the first case and the first death in the State occurred in February 29 and March 24, 2020, and the first of a series of 10 serological surveys (collectively referred to as the EPICOVID19-RS study(4)) in the State was caried out in April 11-13, 2020(5). Although a series of surveys starting so early on is useful for accurately tracking the progression of the epidemic, tests then available that were suitable for population-based studies had only been tested in recently infected individuals. In the EPICOVID19-RS study, a rapid point-of-care test was used. Prior to the 1^st^ survey, available validation studies indicated the test’s performance was sufficiently high for population-based studies(6). However, future validation studies detected that the sensitivity of this test decreases over time(7), an assessment that was only possible after some time since the beginning of the pandemic. In the 9^th^ survey, we used an enzyme-linked immunosorbent assay (ELISA), which has higher sensitivity than the rapid test that does not decrease over time^7-8^. Changing tests between surveys hampers comparing different surveys, thus requiring analytical procedures to properly utilize previous estimates based on the rapid test in combination with the ELISA test to obtain a coherent picture of the pandemic in the state.

External data are extremely useful to furnish procedures to correct biased estimates. The more data the lower the need for modelling assumptions. Typically, such data stems from official statistics, such as registered cases, deaths, hospitalizations and other indicators. However, such data have their own limitations, including dependence of testing policies and protocols, fatality rate, change in treatment protocols, incompleteness and other factors that may change both over time and among different locations.

Obtaining a coherent and likely more accurate temporal trend of how the epidemic in a given location using real data is therefore a challenging task, requiring simultaneously and coherently using different and imperfect sources of data, including official statistics, different diagnostic tests and information from validation studies. This paper aims at obtaining the cumulative prevalence of infection in Rio Grande do Sul from a series of nine population-based serological surveys using a test with temporal variation in sensitivity.

## 2. METHODS

### 2.1. Theoretical justification for the correction procedure

Let 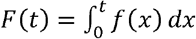 denote the cumulative number of infections at time *t*, where *f* (*x*) is a non-zero function for the infection incidence (in number of new cases) and *t* = 1 is the day of the first infection, so that *F* (1) ≥ 1 and *F*(*t*) = 1 for some *t* ∈ (0,1]. Let *g*(*t*)^*^) ∈[0,1] denote a function for the proportion of infections detectable *t*^*^ days after occurring. Therefore, 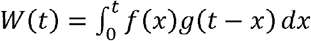 represents the cumulative number of infections detectable at time *t*. We assume the population size *N* is constant throughout the pandemic. In this case,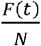 and 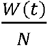 respectively denote the true and apparent (detectable) cumulative prevalence of infections at time *t*.

From the notation above, the proportion of infections that are still detectable (i.e., the sensitivity) in day *t* is:

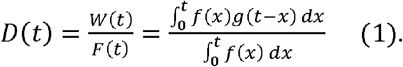

Equation 1 can be interpreted as a weighted average of the proportion of detectable infections, where the weights correspond to the number of infections that occurred in each short interval, [*t, t* + *dt*], where *dt* is a very small positive number.

Let (*h*(*t*) = *cf* (*t*) ⇒ *H* (*t*) = *cF* (*t*), where *c* is an unknown positive constant. *h*(*t*) can denote, for example, the number of deaths, assuming that fatality is constant over time. In this case, *t* represents the time when the infection happened, so that *h*(*t*) denote the number of new cases which ended up dying. If *h*(*t*) denotes the number of deaths in time *t*, then the appropriate representation would be *h*(*t* − *k*) = *cf* (*t*), where *k* denotes the number of days between infection and death.

Substituting *h*(*t*) for *f*(*t*) in equation 1 yields:

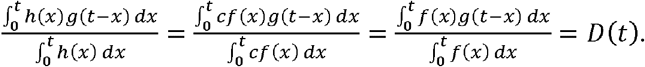

That is, the proportion of infections that are still detectable *t*^*^ days after occurring can also be obtained using any indicator of the temporal distribution of infections, assuming that such indicator is a scaled version of *F*(*t*).

If both that *g* (*t*^*^) and some function *H*(*t*) are known, *g* (*t*^*^) can used to account for the decay in sensitivity over time, and *H* (*t*) can used to account for the temporal distribution of the pandemic. However, in practice, typically only 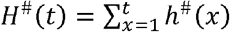, where *h*^#^ (*x*) is the number of deaths that occurred in a given day, is known. From this definition, 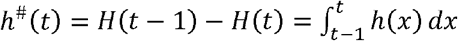 and *H*^#^ (*t*) = *H*(*t*) for *t* ∈ ℕ^*^. Since the exact time of the events within a day are unknown, an approximation can be made by assigning *t*^*^ = *t*^*s*^ - *t* to all events that happened in day *t*, where *t*^*s*^ denotes the date when individuals were tested. This is a good approximation because the *g* (*t*^*^) is unlikely to substantially change within a day – that is, *g* (*t*^*^) ≈ *g* (*t*^*^ + 1).

From the above, *D*(*t*) and *F*(*t*) can be approximated as follows:

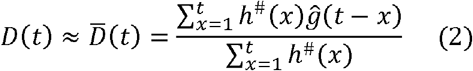

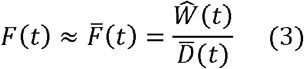

Similar to *D*(*t*) in equation 1, 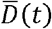 in equation 2 can be interpreted as a weighted average of the proportion of detectable infections, where the weights correspond to the number of infections that occurred in each day.

In practice, *h*^#^ (*x*) can be obtained from official statistics. However, the assumption that is constant over time is required for equation 2 to yield a good approximation of *F*(*t*). If this is not the case, then *c* should be replaced with *c*(*t*). Moreover, it is unlikely that *g* (*t*^*^) is known in practice. However, it can be estimated, for example, in validation studies. Therefore, *ĝ* (*t*^*^) is included in equation 2 instead of *g* (*t*^*^). Finally, equation 3 includes 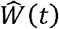, which is the estimated apparent cumulative number of infections estimated using a test with sensitivity that decreases over time, according to *g* (*t*^*^).

### 2.2. Simulation study

For the simulations, 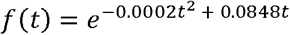, thus yielding ∼1 million individuals who got infected at some point during a one-year period. The population size was 11 million individuals. Fatality rate was 1% and constant over time. These numbers are roughly of comparable magnitude to the first year of the epidemic in Rio Grande do Sul.

Individual *i* (where *i* denotes a generic individual who was infected at some point) was detectable by the Wondfo test at time *t* if 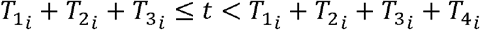, where:

- *T*_1_ is the infection date (defined as described above).
- 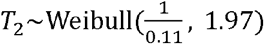 is the incubation time^9-10^.
- *T*_3_ ∼N (13.3, 32.49) is the time from symptom onset to seroconversion(11).
- *T*_4_ ∼ Weibull (68.383320,1.183745) is the time from seroconversion to seroreversion(12).
- *T*_5_ is the time from symptom onset to diagnosis. This was estimated using official statistics (see below for a description of these data) as the time difference between the dates of diagnosis and of symptom onset. For the present analyses, values greater than 21 days were discarded.

Regarding fatality, 1% of the infected individuals were randomly selected as those who died. To define when individuals died, we defined the following transition times:

- *T*_6_ is the time from diagnosis to death. This was estimated using official statistics (see below for a description of these data) as the time difference between the dates of death and of diagnosis.

Among individuals who died in the simulation study, values for *T*_5_-*T*_6_ were generated by sampling from their empirical joint distribution obtained from official statistics, restricting to those diagnosed with RT-PCR and died. For individuals who did not die in the simulation study *T*_5_values were generated by sampling from official statistics, restricting to those diagnosed with RT-PCR and did not die. Therefore, this sampling scheme allows for violations of the assumption that such times are similar between these subgroups. This may introduce some bias in the corrected estimates, and the simulation study will help to quantify how much this bias is likely to affect empirical analysis.

If individual is one of those who died, if 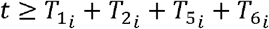, then the *i*th individual was deceased at time *t*. Of note, all times were rounded to the nearest integer, and negative values in *T*_3_ were replaced with zero.

In the simulations, the population size decreases over time as infected individuals die. Deaths due to other causes were not considered because we assumed that, aside from the epidemic, the population size is constant over time. The parameter of interest is 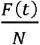,were *N* is assumed to be constant over time. Therefore, this is another source of (likely slight bias) in the estimates, and such bias will be quantified in the simulations.

To generate *g* (*t*^*^), a hypothetical validation study involving 20 million individuals, all enrolled in the day they were infected and each followed up for 400 days, was simulated by independently sampling 20 million values for *T*_2_-*T*_4_ from their respective distributions. Therefore, if 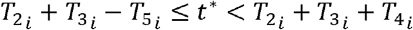, then individual *i* is detectable *t*^*^ days after diagnosis. *g* (*t*^*^) was then defined as the proportion of individuals detectable *t*^*^ after diagnosis. Such large validation study essentially implies that there is negligible error due to sampling variability in *ĝ* (*t*^*^), such that *ĝ* (*t*^*^) ≈ (*t*^*^).

We simulated a scenario of 4 population based serological surveys conducted at days *t* =60, *t* =120 and *t* =180 and *t* =240. For each survey, 5000 individuals were randomly sampled from the population. A total of 1000 datasets were simulated by sampling *T*_2_-*T*_6_ from their respective distributions and randomly allocating these values to the dataset of infected individuals.

The correction procedure was implemented as an iterated procedure. In each dataset, 2000 iterations of the correction procedure for each survey were applied. In each iteration, *H*^#^ (*t*) values for *T*_2_ were sampled from its distribution. These were subtracted from the date of symptom onset, which is assumed to be known for each death (as in our empirical application). This yields a distribution of when individuals who died were infected, which is a valid indicator of the relative temporal distribution of all infections under the assumption that fatality rate is constant over time. This relative temporal distribution, in combination with the sensitivity function *g* (*t*^*^), was then used to calculate 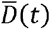 and 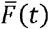.

### 2.3. Data sources

#### 2.3.1. EPCOVID19-RS study

The EPICOVID19-RS study is a series of 10 large, population-based surveys aiming at tracking the COVID-19 pandemic in Rio Grande do Sul. For this paper, only surveys 1 to 9 will be used. Data collection for the 1^st^ and 9^th^ surveys occurred in April 11-13 2020 and February 5-7 2021, respectively. The study involves nine sentinel cities – that is, the largest city from each one of the eight intermediate regions of the state as defined by the Brazilian Institute of Geography and Statistics (IBGE). The exception was the metropolitan region, for which two cities (the state capital and an additional city) were selected. In each city, 50 census tracts (delimited areas with approximately 300 households each in urban area, also defined by the IBGE) were selected with probability proportional to size and ensuring geographical representativity. In each tract, 10 households were systematically selected. Between surveys, different households were selected within the same tracts. One individual per households was randomly selected for interview and testing using the rapid point-of-care lateral-flow WONDFO SARS-CoV-2 Antibody Test (Wondfo Biotech Co., Guangzhou, China), which assesses both IgG and IgM antibodies against SARS-CoV-2. More details on the study protocol are available elsewhere(4). For the present analysis, individuals with inconclusive test results were excluded.

#### 2.3.2. Official statistics data

Official statistics data were collected from a dashboard by the Health Secretariat of Rio Grande do Sul website (https://covid.saude.rs.gov.br/) for the period between February 29, 2020 (the day of the first confirmed case in the State) and February 28, 2021. All 497 city-level health secretariats in Rio Grande do Sul provide information daily on several indicators relevant for monitoring the pandemic in the state, and this information is publicly available per day and per city. We used the daily absolute number of new deaths attributed to COVID-19 as an indicator of the relative temporal distribution of infections in the State. As discussed above, this is only valid under the assumption that fatality is constant over time. We also obtained the daily absolute number of confirmed infections to calculate the cumulative prevalence of confirmed infections for comparison with our corrected estimates.

The cities included in the survey have different population sizes, but all received the same weight in the study (see below for a justification for this). Therefore, weights for each city were generated for the official statistics dataset as the multiplicative inverse of the number of deaths in the corresponding city. This ensures that the distribution of deaths in each city has the same weight in the overall distribution of deaths. A similar weighting scheme was used when calculating the cumulative prevalence of confirmed infections.

### 2.4. Statistical analysis

#### 2.4.1. General description

The goal of the analysis is to combine different data sources to obtain a plausible temporal description of the cumulative prevalence of SARS-CoV-2 infections in Rio Grande do Sul. The main difficulty is that, in the first eight EPICOVID19-RS surveys, only the Wondfo test was used, while both Wondfo and ELISA tests were used. The difficulty stems from the fact that, according to validation studies, the sensitivity of the Wondfo test decreases over time(7). Therefore, any Wondfo seroprevalence estimate after the very beginning of the pandemic is likely to be an underestimate in comparison to the ELISA test.

It is in principle possible to use the results from the validation study to account for this temporal decay in sensitivity, so that sensitivity is assumed to be smaller in later surveys based on some indicator of the temporal distribution of the epidemic, and therefore correction procedures will have a stronger impact on estimates from later surveys. In the 9^th^ survey, the Wondfo seroprevalence was 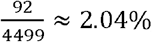, while the ELISA seroprevalence was 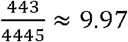.This difference in estimated seroprevalence is not compatible with our validation study, suggesting that sensitivity of the Wondfo test in the field was lower than in the validation study (possibly to the lower prevalence of highly symptomatic and/or severe infection cases in the field, which could influence seroconversion and duration of antibodies over time)(13).

Therefore, the analysis requires two main steps:

a. Calibrating the results from the validation study so that the sensitivity it provides for the 9^th^ survey corresponds to the observed sensitivity, itself estimated by comparting the Wondfo and ELISA estimates in the 9^th^ survey (this simplified procedure for estimating sensitivity is justified below).
b. Use the indicator and the calibrated temporal decay in sensitivity to obtain corrected estimates for surveys 1-8. These corrected estimates can be interpreted as the expected estimates had ELISA been used.

We now describe each one of these steps in detail.

#### 2.4.2. Calibrating the temporal decay in sensitivity

The validation study (details on study design are provided in the Supplement) had data on the date of diagnosis by RT-PCR and of testing by the Wondfo test (thus yielding the time between diagnosis and testing), but not of symptom onset. Therefore, the validation study was re-analyzed in each iteration of the correction procedure. In each iteration:

- The validation study data was resampled without replacement. This allows incorporating sampling variability in the validation study itself as in a non-parametric bootstrap procedure.
- *T*_2_ and *T*_5_ values were generated and added to the time between diagnosis and testing in the resampled dataset to generate *t*^*^.
- A model for the change in sensitivity over time was then fitted by an automated model selection procedure (see the Supplement for details).
- Since the validation study lacked data on sensitivity of the Wondfo test for recent infections, extrapolation for the smallest value of *t*^*^ in the dataset to *t*^*^ = 0 was required. However, the observed data does not capture the increase in sensitivity during the first days following infection. This gap in the range of *t*^*^ was imputed by assuming that sensitivity from *t*^*^ = 0 (assumed to be 1%) to the smallest value of *t*^*^ (using the predicted sensitivity from the model) in the given iteration has a logit-linear relationship.

The process above generated (in each iteration) *ĝ* ′ (*t*^*^), which is the function describing how sensitivity decreased over time in the validation study. As discussed above, the observed differences in seroprevalence estimates from the Wondfo and ELISA tests in the 9^th^ survey were not compatible with our validation study. To account for this difference, we calibrated *ĝ*′ (*t*^*^) into *ĝ* (*t*^*^) = *r*[*ĝ*′ (*t*^*^)], where 0< *r* < 1.

The calibration procedure was performed by finding the value of *r* using a one dimensional optimization algorithm such that 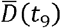 (where *t*_9_ is the date of the 9^th^ survey) is virtually identical to 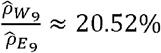 (where 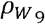 and 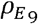 respectively denote the seroprevalence as measured by the Wondfo and ELISA tests in the 9^th^ survey), which is an estimate of the sensitivity of the Wondfo test in the 9^th^ survey. This process ensures that 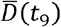 is (virtually) identical to the sensitivity in the last survey, thus making *ĝ* (*t*^*^) and field estimates of sensitivity more compatible.

Importantly, this calibration procedure is arbitrary because many other transformations of *ĝ*′ (*t*^*^)could be used. We decided to simply scale *ĝ*′ (*t*^*^) by a multiplicative constant because this is a simple procedure that is rank preserving, therefore retaining the overall trends of the original function. However, the calibration procedure is entirely numerical, thus being agnostic to biological considerations. Nevertheless, the calibration does return a sensitivity curve that is at least compatible to field estimates, which is the goal of the calibration procedure.

#### 2.4.3. Correcting the Wondfo estimates from surveys 1-8

As described above, the temporal distribution of deaths was used as an indicator of the temporal distribution of the epidemic. However, because the official statistics dataset does not have data when infection occurred (which is indeed virtually impossible to ascertain), the correction procedure was implemented as an iterated process. In each iteration, *T*_2_ values were generated and subtracted from the date at symptom onset (available in the official statistics dataset), thus yielding the infection date *t*.

To calculate corrected cumulative prevalence estimates (denoted by 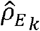) in surveys 1-8, sensitivity (denoted by 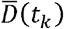) was calculated as the weighted average of *ĝ* (*t*^*^) over the period from February 29, 2020 to the given survey, where the weight that each date receives is the sum of the weights (described in section 2.3.2) of all deaths with infection occurring in the given date. 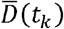 was then applied to the following formula: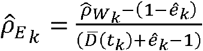, (14) where 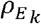 and 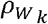 denote the seroprevalence as measured by the ELISA and Wondfo tests (respectively) in the *k*th survey; and *ê*_*k*_ is the specificity of the Wondfo test in the th survey. We assumed *ê*_*k*_= 1 in all surveys, so the formula simplifies to 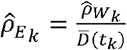.

The strategy outlined above assumes that specificity of the Wondfo test is 100%. Although this is a strong assumption, it can be justified in this context as follows. First, 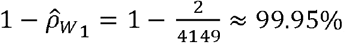, which is a lower bound for the test’s specificity since it was applied in the field. Second, although we have no data on how specificity changes over time, it is likely to increase because the sensitivity decreases. Therefore, *ê*_*k*_ = 1 is likely a good approximation in this situation, which justifies estimating 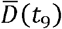 as 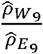, simplifies the formula for calculating 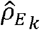 and prevents it from yielding negative values. Although the formula could still yield 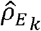 values larger than 100%, this cannot happen in the current application because 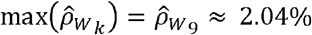 and 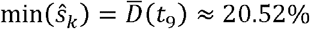, and 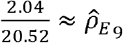. We also performed a subset of the analysis using a likelihood-based approach (described in detail elsewhere(15)) and obtained very similar results.

Confidence intervals of the corrected estimates for surveys 1 to 8 incorporated the different sources of uncertainty in a bootstrap procedure (see Supplement for details). All analyses were performed using R 4.0.2.

## 3. RESULTS

Table 1 displays the results of the simulation study. In comparison to the apparent prevalence, corrected prevalence estimates were on average much closer to the true prevalence. Corrected estimates were slightly biased upwards, but both bias and absolute deviation (that is, the average absolute distance from the true prevalence) were of small magnitude.

**Table 1.** **Performance of the correction procedure in the simulation study (averaged over 1000 datasets).**

| *t* | True<br>prevalence<sup>a</sup> | Apparent<br>prevalence<sup>b</sup> | Corrected values |  |  |
| --- | --- | --- | --- | --- | --- |
|  |  |  | Prevalence<sup>c</sup> | Bias<sup>d</sup> | Absolute<br>bias |
| 60 | 0.011% | 0.002% | 0.014% | 0.003 pp | 0.022 pp |
| 120 | 0.300% | 0.093% | 0.307% | 0.007 pp | 0.114 pp |
| 180 | 2.383% | 0.988% | 2.408% | 0.025 pp | 0.274 pp |
| 240 | 6.503% | 3.051% | 6.518% | 0.015 pp | 0.416 pp |

<sup>a</sup>True cumulative prevalence of infections.
<sup>b</sup>Cummulative prevalence of infections estimated using a test with decaying sensitivity.
<sup>c</sup>Cummulative prevalence of infections estimated by the correction procedure.
<sup>d</sup>Difference between true and corrected prevalence.
pp: Percent points.

Figure 1 shows the cumulative prevalence of infections. The solid line represents the cumulative prevalence of confirmed infections. Estimates from the Wondfo test were not only substantially smaller than corrected estimates (especially for later surveys), but also presented a different overall pattern that would not resemble the remaining results even upon scaling. Moreover, Wondfo estimates were implausible when compared to the cumulative prevalence of infection according to official cases. Up to the 8^th^ survey, the Wondfo and official cumulative prevalence estimates were similar in the additive scale, but in the 9^th^ survey the Wondfo estimate was substantially smaller than the estimate from official statistics, which is a lower bound of the true cumulative prevalence. Table 2 shows the seroprevalence estimates for each survey of the EPICOVID19-RS study using the different methods.

**Table 2.** **Seroprevalence (%) estimates (95% confidence intervals) in Rio Grande do Sul for each survey.**

| Survey | Wondfo |  | ELISA |  |
| --- | --- | --- | --- | --- |
|  | Method | Values | Method | Values |
| 1 | Measured | 0.05 (0.01-0.17) | Modelled | 0.26 (0.03-1.00) |
| 2 | Measured | 0.13 (0.05-0.29) | Modelled | 0.61 (0.19-1.45) |
| 3 | Measured | 0.22 (0.11-0.41) | Modelled | 0.94 (0.39-1.90) |
| 4 | Measured | 0.18 (0.08-0.35) | Modelled | 0.74 (0.28-1.58) |
| 5 | Measured | 0.47 (0.28-0.72) | Modelled | 2.12 (1.12-3.62) |
| 6 | Measured | 0.96 (0.70-1.27) | Modelled | 4.15 (2.55-6.36) |
| 7 | Measured | 1.22 (0.91-1.60) | Modelled | 4.82 (3.09-7.12) |
| 8 | Measured | 1.38 (1.05-1.77) | Modelled | 5.15 (3.40-7.43) |
| 9 | Measured | 2.04 (1.64-2.52) | Measured | 9.97 (9.03-10.96) |

**Figure 1.**
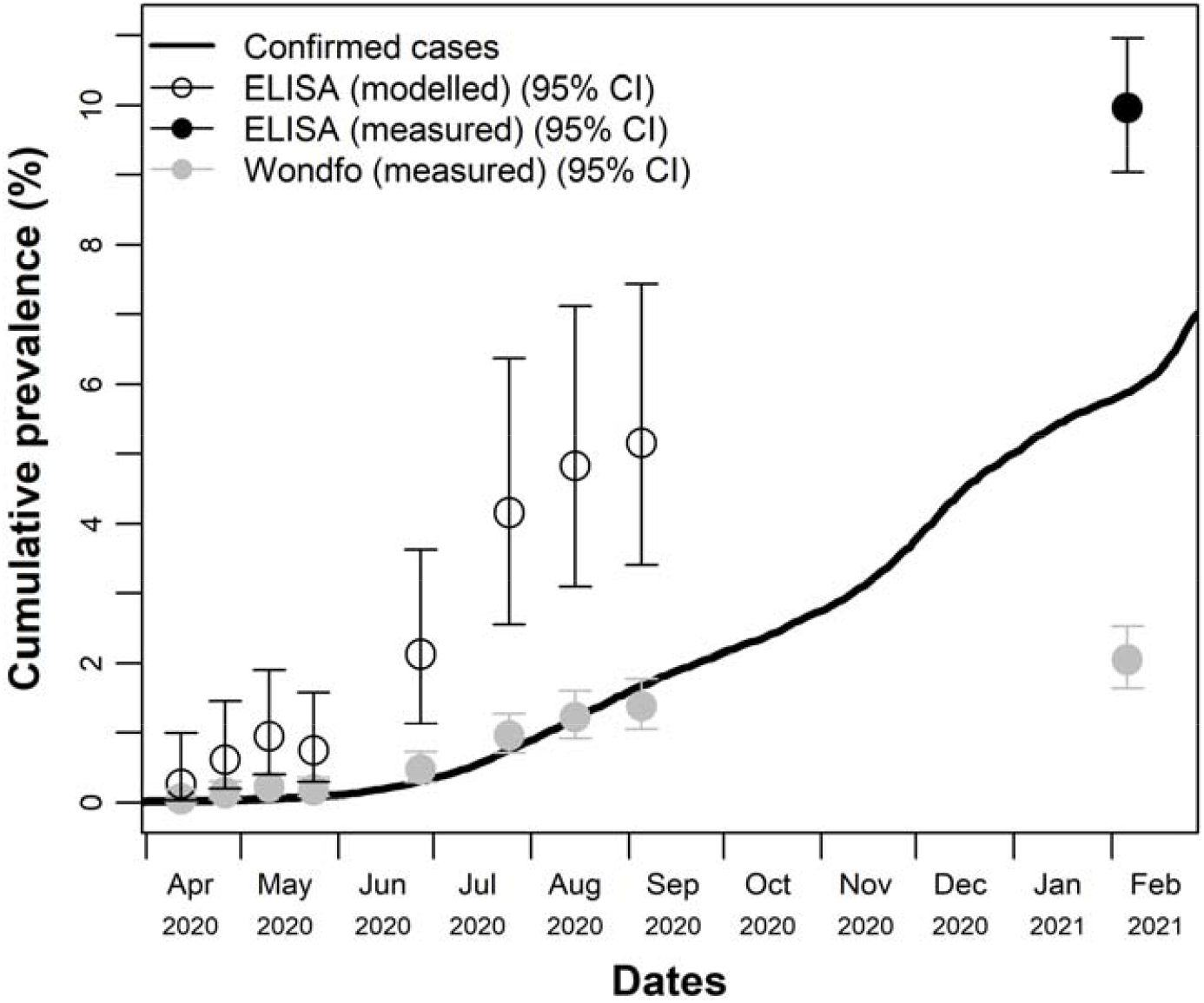
**Cumulative prevalence of SARS-CoV-2 infection (according to the ELISA test) in Rio Grande do Sul (Southern Brazil) from February 29, 2020 to February 28, 2021.**

## 4. DISCUSSION

In this work, we combined data from official statistics, a series of nine population-based serological surveys and a validation study to estimate the cumulative prevalence of SARS-CoV-2 infection in the State of Rio Grande do Sul (Southern Brazil) by adjusting estimates from a rapid test into the expected result had the ELISA test been used instead. Corrected estimates are likely more plausible than estimates using the rapid test, especially when compared to confirmed cases from official statistics. The corrected result indicated that, as of late February 2021, cumulative prevalence of confirmed cases corresponds to about half of all cases detected by ELISA.

Describing the temporal evolution of the epidemic is important for several reasons. First, the true number of infections is required for calculating several important quantities important for policy making, including fatality and hospitalization rates(16). Second, an accurate estimate of the temporal distribution allows better calculation of quantities required for projections using models of infection dynamics, such as the basic reproduction number and how it changes over time, especially after specific events such as lockdown measures or social events that may lead to overcrowding. Projections from disease dynamics models early in the pandemic were highly influential and were one of the few sources of information then available. Although important and useful, it must be noted that most early projects were necessarily based on preliminary estimates from other diseases or from small, non-representative studies. Moreover, it is difficult to a *priori* identify and incorporate all possible complex factors that may influence the temporal evolution of the epidemic, many of which are local and difficult to measure. One year after the first confirmed case in Rio Grande do Sul, there is substantial local data available from official statistics and the EPICOVID19-RS study, thus allowing comparing initial projections to the real figures to refine future projection models, which can now use local, real-data estimates.

The study has considerable strengths. First, the EPICOVID19-RS study is a unique resource that includes nine population-based seroprevalence surveys covered a period of about 10 months and begun shortly after the first confirmed case in the State. Second, the distribution of all but one of the transition times was based on local date. Third, the fact that both the rapid and the ELISA tests were used in the 9^th^ survey allowed us to calibrate the sensitivity function with a result obtained in the field. Fourth, corrected results were more plausible than uncorrected estimates, which underestimated even the cumulative prevalence of confirmed cases after the 8^th^ survey.

An important limitation of the study is the need of many assumptions throughout the correction process, including: fatality ratio is constant over time; and the sensitivity function estimated in the validation study (which was enriched for symptomatic cases) is applicable to the field (which includes the general population) after a calibration procedure. It is not possible to empirically verify the assumption of constant fatality over time without making additional assumptions that may lead to circular reasoning. Regarding the sensitivity function, its validity can only be assessed by repeatedly applying the rapid test over time to individuals sampled from the general population who had been diagnosed with known date. Establishing such cohort would be logistically difficult and time consuming, requiring large initial sample sizes to identify enough infected individuals. It should also be mentioned that the ELISA test is itself not perfect. Indeed, there is evidence indicating that some individuals do not seroconvert, and this may be associated with disease severity(13). Therefore, estimates presented here must be interpreted as the cumulative prevalence of positive ELISA tests rather than of true infections.

This study is not a definitive guide, but rather an example of the usefulness of integrating different sources to correct estimates from an imperfect test to obtain a more plausible temporal trend of the COVID-19 epidemic. Although the study demonstrates the practical importance of analytical procedures, it also highlights the critical importance of good data, including population-based estimates from initiatives such as the EPICOVID19-RS study, which ideally should be performed using appropriate tests; extensive validation studies evaluating how different factors (such as time since diagnosis) can affect the test’s performance; and good quality, transparent and freely accessible official statistics. Estimates from large and populationally-representative data sources with sufficiently fine temporal is the best way to track the progression of the pandemic and to substantiate policy making, and therefore obtaining such data should be supported by funders and governments.

## Data Availability

The data and code is available upon reasonable request.

