## Supplementary File for "Combining serological assays and official statistics to describe the trajectory of the COVID-19 pandemic: results from the EPICOVID19-RS study in Rio Grande do Sul (Southern Brazil)"

**SUMMARY**

1. **SUPPLEMENTARY METHODS**
   1.1. **Fitting the function describing the temporal decay in sensitivity of the rapid test**

In November 2020, a validation study aimed at assessing the sensitivity of the Wondfo and ELISA tests was performed in Pelotas (a city in the state of Rio Grande do Sul, Southern Brazil) in 135 individuals with SARS-CoV-2 infection diagnosed by RT-PCR between April and October 2020. For the ELISA test, 124 tests were positive, 3 were negative and 8 were inconclusive. Therefore, sensitivity was $\frac{\text{124}}{\text{127}}$ $\approx$ 97.8%. For the Wondfo test, there were 85 positive tests, 50 negative tests and no inconclusive results. Therefore, sensitivity was $\frac{\text{85}}{\text{135}}$ $\approx$ $\text{63.0\%}$. Importantly, although there was no time trend in the sensitivity of the ELISA test, the sensitivity of the Wondfo test clearly decreased over time (Supplementary Figure 1). More details on this study can be found elsewhere^1^.

As described in the main text (section 2.4.2), the sensitivity function was fitted and calibrated in an iterated procedure, which involves an automated process for model selection. We now describe the steps that lead to the definition of such automated process. These steps were performed using the observed data of the validation study, without the addition of $T_{2}$ and $T_{5}$.

Supplementary Figure 1 shows the sensitivity functions over time estimated using different methods. A simple logistic regression (black line) specification of the form $\mathrm{logit}\left( \hat{E\left[ R|t^{*} \right]} \right)=\hat{\beta}_{0}+\hat{\beta}_{1}t^{*}$ estimated an almost linear time-dependent decay in sensitivity, which clearly does not correspond well to the observed pattern. Additional functions were estimated using different survival models, assuming that time between events has an exponential (golden line), Gamma (blue line), Log-Normal (pink line) or Weibull (orange line) distribution. These models also yielded functions that did not adjust well to the observed data, overestimating sensitivity for small values of $t^{*}$ and underestimating for large values of $t^{*}$.

One of the reasons for the difficulty to fit a good model to the data is the change in the association between sensitivity and time in different time periods. In the intervals $t^{*}\leq$ 40 and $t^{*}\geq$ 80 (approximately), the association is approximately null or at most weakly negative. In the interval 40 $<t^{*}<$ 80, the association is clearly negative. This motivated assessing a second logistic regression specification including a truncation at $t^{*}=$ 60 (the average of 40 and 80) and a quadratic term to (at least partially) capture additional non-linear associations – that is, a model of the form:

$\mathrm{logit}\left( \hat{E\left[ R|t^{*} \right]} \right)=\hat{\gamma}_{0}+\hat{\gamma}_{1}t^{*}+\hat{\gamma}_{2}{t^{*}}^{2}+\hat{\gamma}_{3}\left( I(t^{*}\geq\text{60})\left( t^{*}-\text{60} \right) \right) (1)$,

where $I\left( t^{*}\geq\text{60} \right)=\left\{ \begin{aligned} \text{0} \mathrm{if} t^{*}<\text{60} \\ \text{1} \mathrm{if} t^{*}\geq\text{60} \end{aligned} \right.$.

This model (red line) fitted the data best among all considered models and was therefore selected as the model for $g'\left( t^{*} \right)$. That is:

$\hat{g}'\left( t^{*} \right)=\mathrm{sigmoid}\left[ \hat{\gamma}_{0}+\hat{\gamma}_{1}t^{*}+\hat{\gamma}_{2}{t^{*}}^{2}+\hat{\gamma}_{3}\left( I(t^{*}\geq\text{60})\left( t^{*}-\text{60} \right) \right) \right] (2)$.

Based on these exploratory analyses, we defined a set of 10 logistic regression models to be fitted to the validation study in each iteration, after resampling with replacement and adding $T_{2}$ and $T_{5}$. The rh.s. of these models is provided below:

- Model 1: $\hat{\gamma}_{0}+\hat{\gamma}_{1}t^{*}$
- Model 2:$\hat{\gamma}_{0}+\hat{\gamma}_{1}t^{*}+\hat{\gamma}_{3}\left( I\left( t^{*}\geq\text{50} \right)\left( t^{*}-\text{50} \right) \right)$
- Model 3:$\hat{\gamma}_{0}+\hat{\gamma}_{1}t^{*}+\hat{\gamma}_{3}\left( I(t^{*}\geq\text{60})\left( t^{*}-\text{6}\text{0} \right) \right)$
- Model 4:$\hat{\gamma}_{0}+\hat{\gamma}_{1}t^{*}+\hat{\gamma}_{3}\left( I(t^{*}\geq\text{60})\left( t^{*}-\text{7}\text{0} \right) \right)$
- Model 5:$\hat{\gamma}_{0}+\hat{\gamma}_{1}t^{*}+\hat{\gamma}_{3}\left( I(t^{*}\geq\text{60})\left( t^{*}-\text{8}\text{0} \right) \right)$
- Model 6:$\hat{\gamma}_{0}+\hat{\gamma}_{1}t^{*}+\hat{\gamma}_{2}{t^{*}}^{2}+\hat{\gamma}_{3}{t^{*}}^{3}$
- Model 7:$\hat{\gamma}_{0}+\hat{\gamma}_{1}t^{*}+\hat{\gamma}_{2}{t^{*}}^{2}+\hat{\gamma}_{3}\left( I\left( t^{*}\geq\text{50} \right)\left( t^{*}-\text{50} \right) \right)$
- Model 8:$\hat{\gamma}_{0}+\hat{\gamma}_{1}t^{*}+\hat{\gamma}_{2}{t^{*}}^{2}+\hat{\gamma}_{3}\left( I\left( t^{*}\geq\text{6}\text{0} \right)\left( t^{*}-\text{6}\text{0} \right) \right)$
- Model 9:$\hat{\gamma}_{0}+\hat{\gamma}_{1}t^{*}+\hat{\gamma}_{2}{t^{*}}^{2}+\hat{\gamma}_{3}\left( I\left( t^{*}\geq\text{7}\text{0} \right)\left( t^{*}-\text{7}\text{0} \right) \right)$
- Model 10:$\hat{\gamma}_{0}+\hat{\gamma}_{1}t^{*}+\hat{\gamma}_{2}{t^{*}}^{2}+\hat{\gamma}_{3}\left( I\left( t^{*}\geq\text{8}\text{0} \right)\left( t^{*}-\text{8}\text{0} \right) \right)$

Model 1 was included because it is the simplest model, and virtually always yields a model where sensitivity decreases over time. Models 2-5 are two-parameter models, consisting of simpler versions of the truncated model in equation 1, with varying cut-offs due to the variability introduced by adding $T_{2}$ and $T_{5}$. Models 6-10 are three-parameter models, consisting of a 3^rd^ degree polynomial (model 6) and four truncated models including a quadratic terms (all variations of the model in equation 1).

In each iteration, the 10 models were fitted to the data. The model with the smallest AIC that satisfied the condition $\hat{g}'\left( 50 \right)\leq\hat{g}'\left( 100 \right)\leq\hat{g}'\left( 150 \right)\leq\hat{g}'\left( 200 \right)$ (that is, had an overall pattern of non-increasing sensitivity for 50 $\leq t^{*}\leq200$) was selected. If no model fulfilled these criteria, the iteration was repeated and the process repeated. Importantly, this imposition had little practical effect because model 1 virtually always satisfied that non-increasing condition.

It should be noted that $\hat{g}^{'}\left( t^{*} \right)$ was calculated using RT-PCR as the reference (or “gold-standard”) method. However, in the main text we report correct estimates with reference to the ELISA test. The reason why RT-PCR was used for fitting $\hat{g}^{'}\left( t^{*} \right)$ is that it allowed including all 135 individuals in the analysis, which is useful especially because there were very few individuals for some values of $t^{*}$. Moreover, since the sensitivity of the ELISA test was $\approx$97.8%, a sensitivity function estimated with reference to ELISA would be very similar to $\hat{g}^{'}\left( t^{*} \right)$. Finally, $\hat{g}^{'}\left( t^{*} \right)$ was calibrated (section 2.4.2 in the main text) so that the sensitivity in the 9^th^ survey matches the observed sensitivity of the Wondfo test in relation to the ELISA test.

1.2. **Bootstrap procedure for calculating confidence intervals**

For a given corrected seroprevalence estimate, there is sampling variation in: the Wondfo estimate, denoted by ${\hat{\rho}_{W}}_{k}$; the observed sensitivity in the 9^th^ survey, denoted by $\bar{D}_{9}$; the estimated sensitivity based on $\hat{g}\left( t^{*} \right)$; and $T_{2}$ and $T_{5}$. Since only Wondfo estimates for surveys 1-8 were corrected, $\bar{D}_{9}$ was estimated in survey 9 and $\hat{g}^{'}\left( t^{*} \right)$ was fitted in an independent validation study, these parameters were assumed to be independent.

To account for the sampling design, all survey data analyses were performed using the functionality provided by the “survey” package^3,4^. Because municipalities were selected *a priori* and census tracts were sampled within each municipality, census tracts were treated as principal sampling units and municipalities as strata. Importantly, the fact that census tracts were sampled with probability proportionate to size and a fixed number of households was sampled in each tract, this is a self-weighted design. No weighting for population size (or any other type of weighting) was performed. Variance was estimated analytically using Taylor series linearization estimation (this procedure is described in detail elsewhere^3^).

To incorporate these three sources of uncertainty and the sampling design in the confidence interval for ${\hat{\rho}_{E}}_{k}$ (the estimated seroprevalence in survey $k$ had ELISA been used), we used bootstrap, as described below. Within each iteration of the bootstrap procedure, values for $T_{2}$ were generated and applied to the official statistics dataset of deaths; and values for $T_{2}$ and $T_{5}$ were generated and applied to the resampled validation study dataset. All these values were generated independently, sampling from their respective univariate distributions. A total of 50,000 iterations were performed, each indexed by $j\in\left\{ \text{1,…,50,000} \right\}$. All analyses were performed using R 4.0.2^5^.

1.2.1. ***Generating the empirical sampling distribution of*** ${\boldsymbol{\rho}_{\boldsymbol{W}}}_{\boldsymbol{k}}$

This was performed in the following steps:

- Calculate design-adjusted standard errors for ${\hat{\rho}_{W}}_{k}$ (denoted as ${\sigma_{W}}_{k}$) by first fitting an intercept-only logistic regression model having the test result as the dependent variable. We then used this model to estimate ${\sigma_{W}}_{k}$ using the “predict” function.
- Calculate the effective sample size ($N_{k}^{*}$) – i.e., the sample size that a study using simple random sampling would be expected to have so that the standard error of ${\hat{\rho}_{W}}_{k}$ equals ${\sigma_{W}}_{k}$. This was calculated as $N_{k}^{*}=\min\left[ N_{k},\frac{{\hat{\rho}_{W}}_{k}\left( 1-{\hat{\rho}_{W}}_{k} \right)}{{\sigma_{W}^{2}}_{k}} \right]$, where $N_{k}$ is the actual sample size. $N_{k}^{*}$ was rounded to the nearest integer.
- Generate the empirical sampling distribution of ${\rho_{W}}_{k}$ as ${{\hat{\rho}_{W}}_{k}}_{j}\sim\frac{B\left( N_{k}^{*},{\hat{\rho}_{W}}_{k} \right)}{N_{k}^{*}}$. Importantly, by using $N_{k}^{*}$ instead of $N_{k}$ in this step, the variance of the empirical sampling distribution is ${\sigma_{W}^{2}}_{k}$, thus accounting for the sampling design.
  1.2.2. ***Generating the empirical sampling distribution of*** ${\bar{\boldsymbol{D}}}_{\boldsymbol{9}}$

This was performed in the following steps:

- Fit a logistic regression model of the form $\mathrm{logit}\left( \hat{E\left[ E|W \right]} \right)=\hat{\beta}_{0}+\hat{\beta}_{1}W$, where $E$ and $W$ respectively denote the ELISA and Wondfo tests results in the 9^th^ survey. Let $\hat{\mu}=\left[ \begin{matrix} \hat{\beta}_{0} \\ \hat{\beta}_{1} \end{matrix} \right]$ denote the coefficient vector and $\mathbf{C}=\left[ \begin{matrix} var(\hat{\beta}_{0}) & cov(\hat{\beta}_{0},\hat{\beta}_{1}) \\ cov(\hat{\beta}_{0},\hat{\beta}_{1}) & var(\hat{\beta}_{1}) \end{matrix} \right]$ denote its variance-covariance matrix.
- Generate the empirical sampling distribution of $\mu$ as $\left( {\hat{\beta}_{0}}_{j},{\hat{\beta}_{1}}_{j} \right)\sim N\left( \hat{\mu},\mathbf{C} \right)$.
- Generate the empirical sampling distribution of $\bar{D}_{9}$ as ${\bar{D}_{9}}_{j}=\left( 1-{{\hat{\rho}_{W}}_{9}}_{j} \right)\mathrm{sigmoid}\left( {\hat{\gamma}_{0}}_{j} \right)+{{\hat{\rho}_{W}}_{9}}_{j}\mathrm{sigmoid}\left( {\hat{\gamma}_{0}}_{j}+{\hat{\gamma}_{1}}_{j} \right)$.
  1.2.3. ***Generating the empirical sampling distribution of*** ${{\hat{\boldsymbol{\rho}}}_{\boldsymbol{E}}}_{\boldsymbol{k}}$ ***and calculating confidence intervals***

This was performed in the following steps:

- Estimate $\hat{g}\left( t^{*} \right)$ as described in section 1.1 above and section 2.4.2 in the main text). Let $\hat{g}_{j}\left( t^{*} \right)$ denote the sensitivity function estimated in the $j$th iteration.
- Generate $h_{j}^{\#}\left( t \right)$ – that is, the distribution of infection dates of cases which ended up dying – by subtracting ${T_{2}}_{j}$ from the date at symptom onset.
- Use $h_{j}^{\#}\left( t \right)$ and $\hat{g}_{j}\left( t^{*} \right)$ to calculate ${\bar{D}_{k}}_{j}$ (the sensitivity in the $k$th survey and $j$th iteration) as shown in equation 2 in the main text.
- Generate the empirical sampling distribution of ${\rho_{E}}_{k}$ as ${{\hat{\rho}_{E}}_{k}}_{j}=\frac{{{\hat{\rho}_{W}}_{k}}_{j}}{{\bar{D}_{k}}_{j}}$ (see section 2.4.3 in the main text for a justification for this correction formula).
- Update the effective sample size ($N_{k}^{'}$) as follows: $N_{k}^{'}=\min\left[ N_{k}^{*},\frac{{\hat{\rho}_{E}}_{k}\left( 1-{\hat{\rho}_{E}}_{k} \right)}{\sigma_{E}^{2}} \right]$, rounded to the nearest integer. $\sigma_{E}^{2}$ is the standard deviation of the empirical sampling distribution of ${\rho_{E}}_{k}$.
- Calculate the effective number of positive tests as $n_{p}^{'}={\hat{\rho}_{E}}_{k}N^{'}$.
- Use $n_{p}^{'}$ and $N^{'}$ to calculate the exact binomial confidence interval. When $n_{p}^{'}$ is not an integer, we opted by not rounding it because, due to the small number of positive tests, any rounding would correspond to a substantial relative change in the prevalence. To overcome this issue, we calculated to confidence intervals: one for the nearest smaller integer (i.e., $\left\lfloor n_{p}^{'} \right\rfloor$) and another for the nearest larger integer (i.e., $\left\lceil n_{p}^{'} \right\rceil$). Let $a_{1}$ and $b_{1}$ respectively denote the lower and upper limits of the confidence interval using $\left\lfloor n_{p}^{'} \right\rfloor$, and $a_{2}$ and $b_{2}$ denote the same for $\left\lceil n_{p}^{'} \right\rceil$. The confidence interval for $n_{p}^{'}$ was then calculated as follows: $a=\sum_{i=1}^{2} a_{i}w_{i}$ and $b=\sum_{i=1}^{2} b_{i}w_{i}$, where $w_{i}$ is the weight that each confidence interval receives, calculated as follows: $w_{1}=1-\left( n_{p}^{'}-\left\lfloor n_{p}^{'} \right\rfloor\right)$ and $w_{2}=\left( n_{p}^{'}-\left\lfloor n_{p}^{'} \right\rfloor\right)$.

2. **SUPPLEMENTARY REFERENCES**

1.	Silveira MF, Mesenburg M, Dellagostin OA, Oliveira NR, Maia MAC, Santos FDS, et al. Time-Dependent Decay of Detectable Antibodies Against SARS-CoV-2: A Comparison of ELISA with Two Batches of a Lateral-Flow Test. SSRN Electron J [Internet]. 2021 Jan 24 [cited 2021 Mar 10]; Available from: https://papers.ssrn.com/abstract=3757411

2.	Brent RP. Algorithms for Minimization Without Derivatives. Mineola, NY, USA: Dover Publications; 2013. 206 p.

3.	Lumley T. Analysis of complex survey samples [Internet]. Vol. 9, Journal of Statistical Software. American Statistical Association; 2004 [cited 2021 Mar 11]. p. 1–19. Available from: https://www.jstatsoft.org/index.php/jss/article/view/v009i08/paper-5.pdf

4.	Lumley T. survey: analysis of complex survey samples. R package version 3.35-1 [Internet]. 2019. Available from: https://cran.r-project.org/package=survey

5.	R Core Team. R: A Language and Environment for Statistical Computing [Internet]. Vienna, Austria; 2020. Available from: https://www.r-project.org/

3. **SUPPLEMENTARY FIGURE**

**Supplementary Figure 1. Observed (in 30-day windows) and estimated sensitivity as a function of time between diagnosis by RT-PCR and Wondfo test using several modelling approaches. The dotted line indicates a time difference of 60 days.**

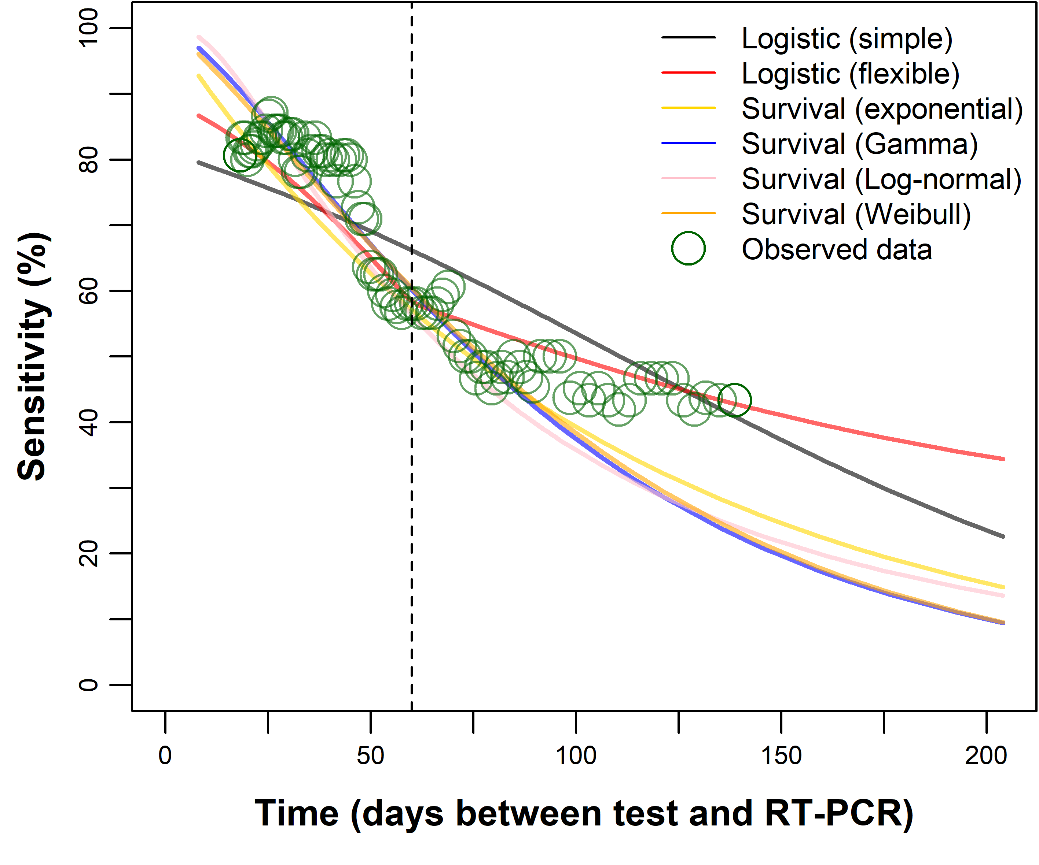
